# Clinical characteristics of 82 death cases with COVID-19

**DOI:** 10.1101/2020.02.26.20028191

**Authors:** Bicheng Zhang, Xiaoyang Zhou, Yanru Qiu, Fan Feng, Jia Feng, Yifan Jia, Hengcheng Zhu, Ke Hu, Jiasheng Liu, Zaiming Liu, Shihong Wang, Yiping Gong, Chenliang Zhou, Ting Zhu, Yanxiang Cheng, Zhichao Liu, Hongping Deng, Fenghua Tao, Yijun Ren, Biheng Cheng, Ling Gao, Xiongfei Wu, Lilei Yu, Zhixin Huang, Zhangfan Mao, Qibin Song, Bo Zhu, Jun Wang

## Abstract

**Background:** A recently developing pneumonia caused by SARS-CoV-2 was originated in Wuhan, China, and has quickly spread across the world. We reported the clinical characteristics of 82 death cases with COVID-19 in a single center.

**Methods:** Clinical data on 82 death cases laboratory-confirmed as SARS-CoV-2 infection were obtained from a Wuhan local hospital’s electronic medical records according to previously designed standardized data collection forms.

**Results:** All patients were local residents of Wuhan, and the great proportion of them were diagnosed as severe illness when admitted. Most of the death cases were male (65.9%). More than half of dead patients were older than 60 years (80.5%) and the median age was 72.5 years. The bulk of death cases had comorbidity (76.8%), including hypertension (56.1%), heart disease (20.7%), diabetes (18.3%), cerebrovascular disease (12.2%), and cancer (7.3%). Respiratory failure remained the leading cause of death (69.5%), following by sepsis syndrome/MOF (28.0%), cardiac failure (14.6%), hemorrhage (6.1%), and renal failure (3.7%). Furthermore, respiratory, cardiac, hemorrhage, hepatic, and renal damage were found in 100%, 89%, 80.5%, 78.0%, and 31.7% of patients, respectively. On the admission, lymphopenia (89.2%), neutrophilia (74.3%), and thrombocytopenia (24.3%) were usually observed. Most patients had a high neutrophil-to-lymphocyte ratio of >5 (94.5%), high systemic immune-inflammation index of >500 (89.2%), increased C-reactive protein level (100%), lactate dehydrogenase (93.2%), and D-dimer (97.1%). A high level of IL-6 (>10 pg/ml) was observed in all detected patients. Median time from initial symptom to death was 15 days (IQR 11-20), and a significant association between aspartate aminotransferase (p=0.002), alanine aminotransferase (p=0.037) and time from initial symptom to death were interestingly observed.

**Conclusion:** Older males with comorbidities are more likely to develop severe disease, even die from SARS-CoV-2 infection. Respiratory failure is the main cause of COVID-19, but either virus itself or cytokine release storm mediated damage to other organ including cardiac, renal, hepatic, and hemorrhage should be taken seriously as well.

**Funding:** No founding.

**Research in context:** *Evidence before this study:* As the seventh member of enveloped RNA coronavirus, severe acute respiratory syndrome coronavirus (SARS-CoV)-2 causes a cluster of severe respiratory disease which is similar to another two fatal coronavirus infection caused by SARS-CoV and Middle Eastern respiratory syndrome coronavirus (MERS-CoV). Through searching PubMed and the China National knowledge infrastructure databases up to February 20, 2020, no published article focusing on hospitalized dead patients was identified.

*Added value of this study:* We conducted a single-center investigation involving 82 hospitalized death patients with COVID-19 and focused on their epidemiological and clinical characteristics. 66 of 82 (80.5%) of patients were older than 60 years and the median age was 72.5 years. The bulk of death cases had comorbidity (76.8%). Respiratory failure remained the leading cause of death, following by sepsis syndrome/MOF, cardiac failure, hemorrhage, and renal failure. Most patients had a high neutrophil-to-lymphocyte ratio, high systemic immune-inflammation index, and increased levels of proinflammatory cytokines.

*Implications of all the available evidence:* SARS-CoV-2 causes a cluster of severe respiratory illness which is similar to another two fatal coronavirus infection caused by SARS-CoV and MERS-CoV. Death is more likely to occur in older male patients with comorbidity. Infected patients might develop acute respiratory distress and respiratory failure which was the leading cause of death, but damages of other organs and systems, including cardiac, hemorrhage, hepatic, and renal also contribute to the death. These damages might be attributable to indirect cytokines storm initiated by immune system and direct attack from SARS-CoV-2 itself.

## Introduction

In December 2019, the first acute respiratory disease caused by severe acute respiratory syndrome coronavirus (SARS-CoV)-2, and recently officially named as Corona Virus Disease 2019 (COVID-19) by World Health Organization (WHO) occurred in Wuhan, China.^1,2^ Person-to-person transmission has been identified through respiratory droplets or likely feces.^3-5^ By February 14, 2020, more than 60,000 confirmed cases and close to 2,000 dead cases have been documented in China, with hundreds of imported patients found in other countries.^4-7^

Generally, the incubation period of COVID-19 was 3 to 7 days. Fever, cough, and fatigue were the most common symptoms.^1^ Approximately 20-30% of cases would develop severe illness, and some need further intervention in intensive care unit (ICU).^8,9^ Organ dysfunction including acute respiratory distress syndrome (ARDS), shock, acute cardiac injury, and acute renal injury, can happen in severe cases with COVID-19.^1,8,9^ It has been reported that critical ill patients were more likely to be older, had underlying diseases, and were more likely to have a symptom of dyspnea.^9^ Oxygen therapy, mechanical ventilation, intravenous antibiotics and antiviral therapy were usually applied in clinical management, but presently there were no effective drugs for improving the clinical outcome of COVID-19, especially for severe cases.^1,8,9^

ARDS, a rapidly progressive disease, is the main cause of death for the patients infected with previously recognized corona virus infection such as SARS-CoV and Middle Eastern respiratory syndrome coronavirus (MERS-CoV).^10,11^ In this context, it was initially considered that lung is the most commonly damaged organ by SARS-CoV-2 infection since human airway epithelia express angiotensin converting enzyme 2 (ACE2), a host cell receptor for SARS-CoV-2 infection.^12,13^ However, increasing clinical cases indicated cardiac, renal and even digestive organ damage in the patients with COVID-19,^9^ which is consistent with the findings that kidney, colon and the other tissues also express ACE2 besides airway epithelia.^14,15^ The above clinical phenomenon and basic research suggest more complicated pathogenesis of COVID-19. Hence, analyzing clinical characteristics of death cases with COVID-19 is urgently needed to improve the outcome of infected patients.

## Methods

### Data collection

We retrospectively collected epidemiological and clinical features of laboratory-confirmed COVID-19 dead patients from January 11, 2020 to February 10, 2020 in the Renmin Hospital, Wuhan University. The confirmed diagnosis of COVID-19 was defined as a positive result by using real-time reverse-transcriptase polymerase-chain-reaction (RT-PCR) detection for routine nasal and pharyngeal swab specimens. This study received approval from the Research Ethics Committee of the Renmin Hospital of Wuhan University, Wuhan, China (approval number: WDRY2020-K038). The Research Ethics Committee waived the requirement informed consent before the study started because of the urgent need to collect epidemiological and clinical data. We analyzed all the data anonymously.

The clinical features, including clinical symptoms, signs, laboratory analyses, radiological findings, treatment, and outcome, were obtained from the hospital’s electronic medical records according to previously designed standardized data collection forms. Laboratory analyses included complete blood count, liver function, renal function, electrolytes test, coagulation function, C-reactive protein, lactate dehydrogenase, myocardial enzymes, procalcitonin, and status of other virus infection. Radiological analyses comprised of X-ray and computed tomography.

The date of onset of symptoms, initial diagnosis of COVID-19, and death were recorded accurately. The incubation period was defined as the time from the contact of transmission origin to the onset of different symptoms and signs. Onset survival time was defined as the period between the onset of different symptoms and signs and the time of death. To increase the accuracy of collected data, two researchers independently reviewed the data collection forms. We also directly communicated with patients or their family members to ascertain the epidemiological and symptom data.

### Inflammation markers

Inflammation markers were calculated using specific parameters of blood tests. Neutrophil-to-lymphocyte ratio (NLR) was calculated by dividing the absolute neutrophil count by the lymphocyte count. Systematic inflammatory index (SII) was defined as platelet count × neutrophil count/lymphocyte count (/μL). Interleukin (IL)-6 was detected using Human Cytokine Standard Assays panel (ET Healthcare, Inc., Shanghai, China) and the Bio-Plex 200 system (Bio-Rad, Hercules, CA, USA) according to the manufacturer’s instructions.

### Statistical analysis

Descriptive analyses were used to determine the patients’ epidemiological and clinical features. Continuous variables were presented as median and interquartile range (IQR), and categorical variables were expressed as the percentages in different categories. The Chi-squared test or Fisher’s exact test was adopted for category variables. The association between the different clinical variables and the time from initial symptom to death was evaluated using Spearman’s rank correlation coefficient. Statistical analyses in this study were performed with use of STATA 15.0 software (Stata Corporation, College Station, TX, USA). A two-sided p value less than 0.05 was considered statistically significant.

## Results

From January 11, 2020 to February 10, 2020, a total of 1,334 patients with a diagnosis of laboratory-confirmed COVID-19 were recorded in the Renmin Hospital, Wuhan University, while 6.2% (82/1334) of patients with this disease were dead. In the same period, the rate of mortality for all causes and non-COVID-19 in this hospital, were 2.3% (162/7119) and 1.4% (80/5785), respectively. The mortality rate of COVID-19 was higher than that of non-COVID-19 (p<0.001).

The epidemiological features and underlying diseases were shown in table 1. All patients were local residents of Wuhan, and only 2 patients acknowledged a contact with the patients confirmed as SARS-CoV-2 infection. All the patients denied a history of contact with wildlife or Huanan seafood market visit. The great proportion of them were diagnosed as severe illness when admitted (77/82). Median incubation time was 7 days (IQR 5.0-10.0). Most of the death cases were male (65.9%), older than 60 years (80.5%) and the median age was 72.5 years (IQR 65.0-80.0). The bulk of death cases had comorbidity (75.6%), including hypertension (56.1%), heart disease (20.7%), diabetes (18.3%), cerebrovascular disease (12.2%), and cancer (7.3%). 30 out of 82 dead patients (30.5%) had 2 or more underlying diseases.

**Table 1.** Clinical features of dead patients with COVID-19

| **Clinical features** |  |
| --- | --- |
| **Age, years** | 72.5 (65.0-80.0) |
| **Age group** |  |
| ≤40 years | 2/82 (2.4) |
| 40-50 years | 4/82 (4.9) |
| 50-60 years | 10/82 (12.2) |
| 60-70 years | 20/82 (24.4) |
| 70-80 years | 26/82 (31.7) |
| >80 years | 20/82 (24.4) |
| **Sex** |  |
| Male | 54/82 (65.9) |
| Female | 28/82 (34.1) |
| **Local residents of Wuhan** | 82/82 (100) |
| **Severe pneumonia** | 77/82 (93.9) |
| **Median incubation time (range), days** | 7/82 (5-10) |
| **Comorbidity** |  |
| All | 62/82 (76.8) |
| Hypertension | 46/82 (56.1) |
| Diabetes | 15/82 (18.3) |
| Chronic obstructive pulmonary disease | 12/82 (14.6) |
| Heart disease | 17/82 (20.7) |
| Cerebrovascular | 10/82 (12.2) |
| Liver disease | 2/82 (2.4) |
| Renal insufficiency | 4/82 (4.9) |
| Infection | 5/82 (6.1) |
| Cancer | 6/82 (7.3) |
| Surgery | 3/82 (3.7) |
| Disease with reduced immunity | 14/82 (17.1) |
| **Number of comorbidity diseases** |  |
| 0 | 19/82 (23.2) |
| 1 | 25/82 (30.5) |
| 2 | 18/82 (22.0) |
| ≥3 | 7/82 (8.5) |

Data are presented as median (IQR), n/N (%), where N represents the total number of patients with COVID-19

We analyzed the causes of mortality of patients with laboratory-confirmed SARS-CoV-2 infection (table 2). Respiratory failure remained the leading cause of death (69.5%), following by sepsis syndrome/multiple organ dysfunction syndrome (MOF) (28.0%), cardiac failure (14.6%), hemorrhage (6.1%), and renal failure (3.7%). Furthermore, respiratory, cardiac, hemorrhage, hepatic, and renal damage were found in 100%, 89%, 80.5%, 78.0%, and 31.7% of patients, respectively. A majority of patients (75.6%) had 3 or more damaged organs or systems following the infection with SARS-CoV-2.

**Table 2.**
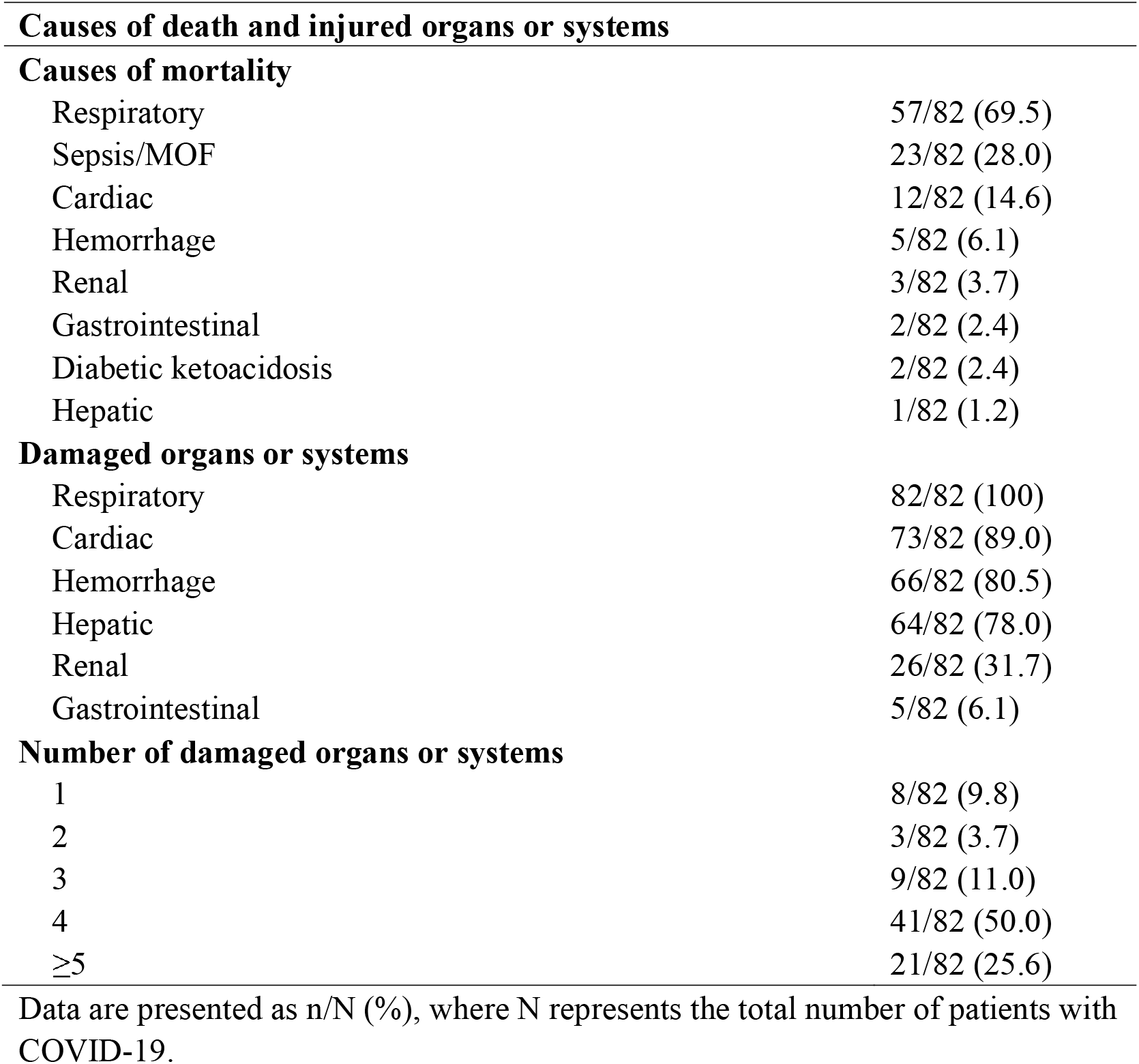
Causes of death of patients with COVID-19

|  |  |
| --- | --- |
| Respiratory | 57/82 (69.5) |
| Sepsis/MOF | 23/82 (28.0) |
| Cardiac | 12/82 (14.6) |
| Hemorrhage | 5/82 (6.1) |
| Renal | 3/82 (3.7) |
| Gastrointestinal | 2/82 (2.4) |
| Diabetic ketoacidosis | 2/82 (2.4) |
| Hepatic | 1/82 (1.2) |
| **Damaged organs or systems** |  |
| Respiratory | 82/82 (100) |
| Cardiac | 73/82 (89.0) |
| Hemorrhage | 66/82 (80.5) |
| Hepatic | 64/82 (78.0) |
| Renal | 26/82 (31.7) |
| Gastrointestinal | 5/82 (6.1) |
| **Number of damaged organs or systems** |  |
| 1 | 8/82 (9.8) |
| 2 | 3/82 (3.7) |
| 3 | 9/82 (11.0) |
| 4 | 41/82 (50.0) |
| ≥5 | 21/82 (25.6) |

Data are presented as n/N (%), where N represents the total number of patients with COVID-19.

As shown in table 3, fever (78.0%), cough (64.6%), and shortness of breath (63.4%) were the main common symptom. Diarrhea was observed in 12.2% of patients. All patients had bilateral involvement of chest radiographs. On the admission, lymphopenia (89.2%), neutrophilia (74.3%), and thrombocytopenia (24.3%) were usually observed. All the patients had increased C-reactive protein level (100%), and most patients had a high NLR >5.0 (94.5%), SII index of >500 (89.2%), lactate dehydrogenase (93.2%), D-dimer (97.1%), cardiac troponin I (86.7%), and procalcitonin (81.2%). Insufficient cell immunity with reduced CD3+ (93.1%), CD8+ (98.3%), CD6+CD56+ (100%) cell count, and high level of circulating IL-6 (100%) were observed in patients.

**Table 3.**
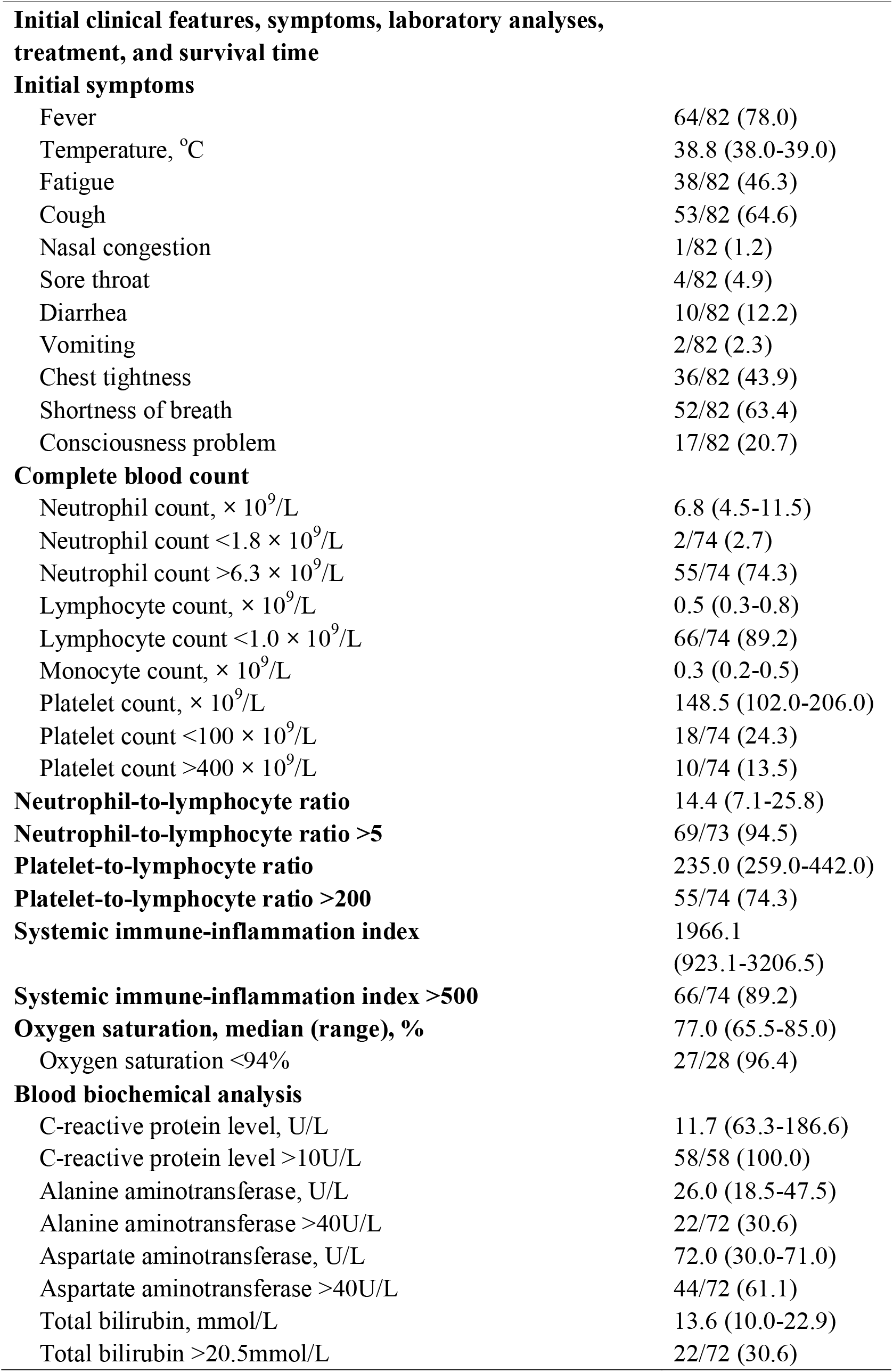
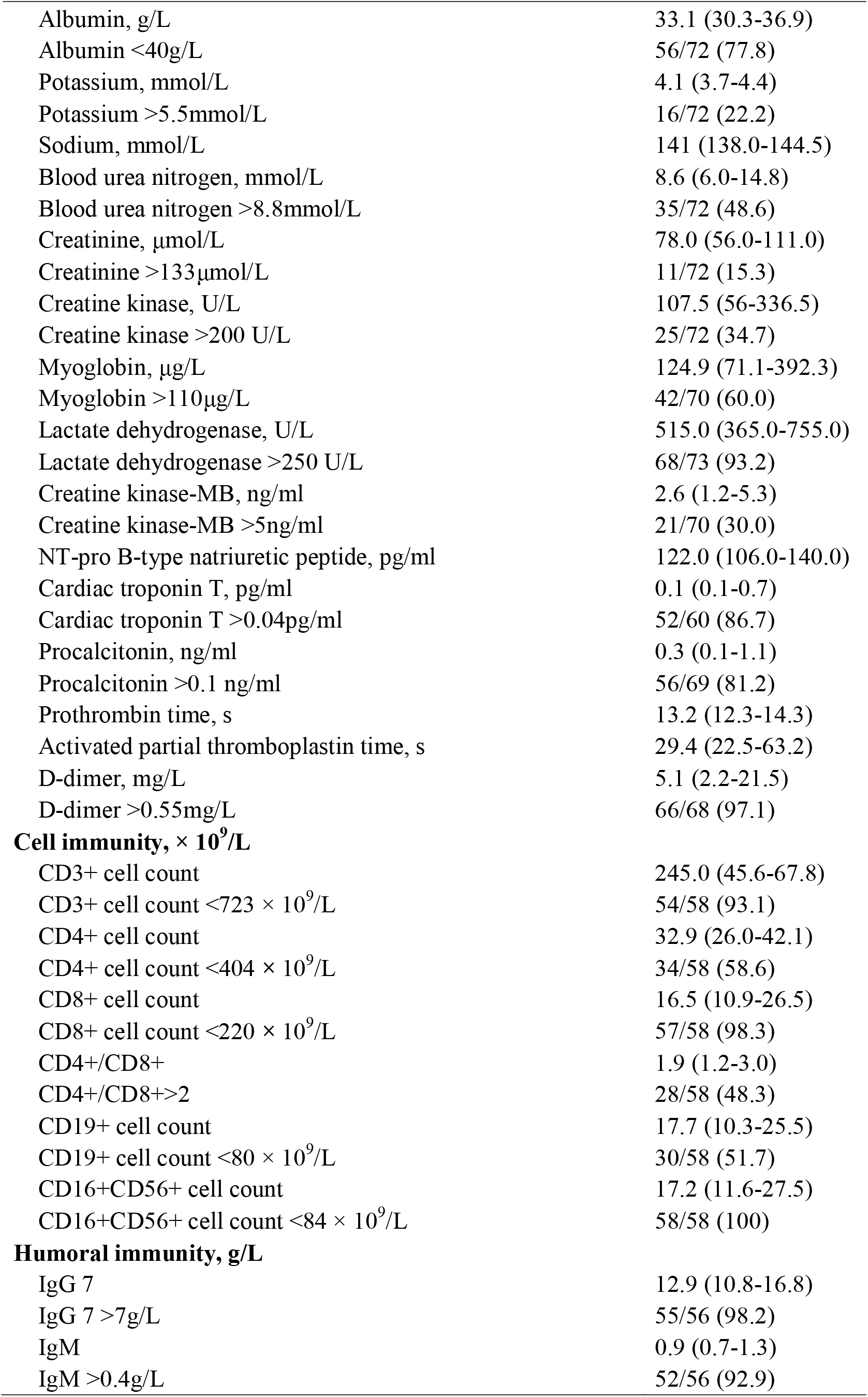
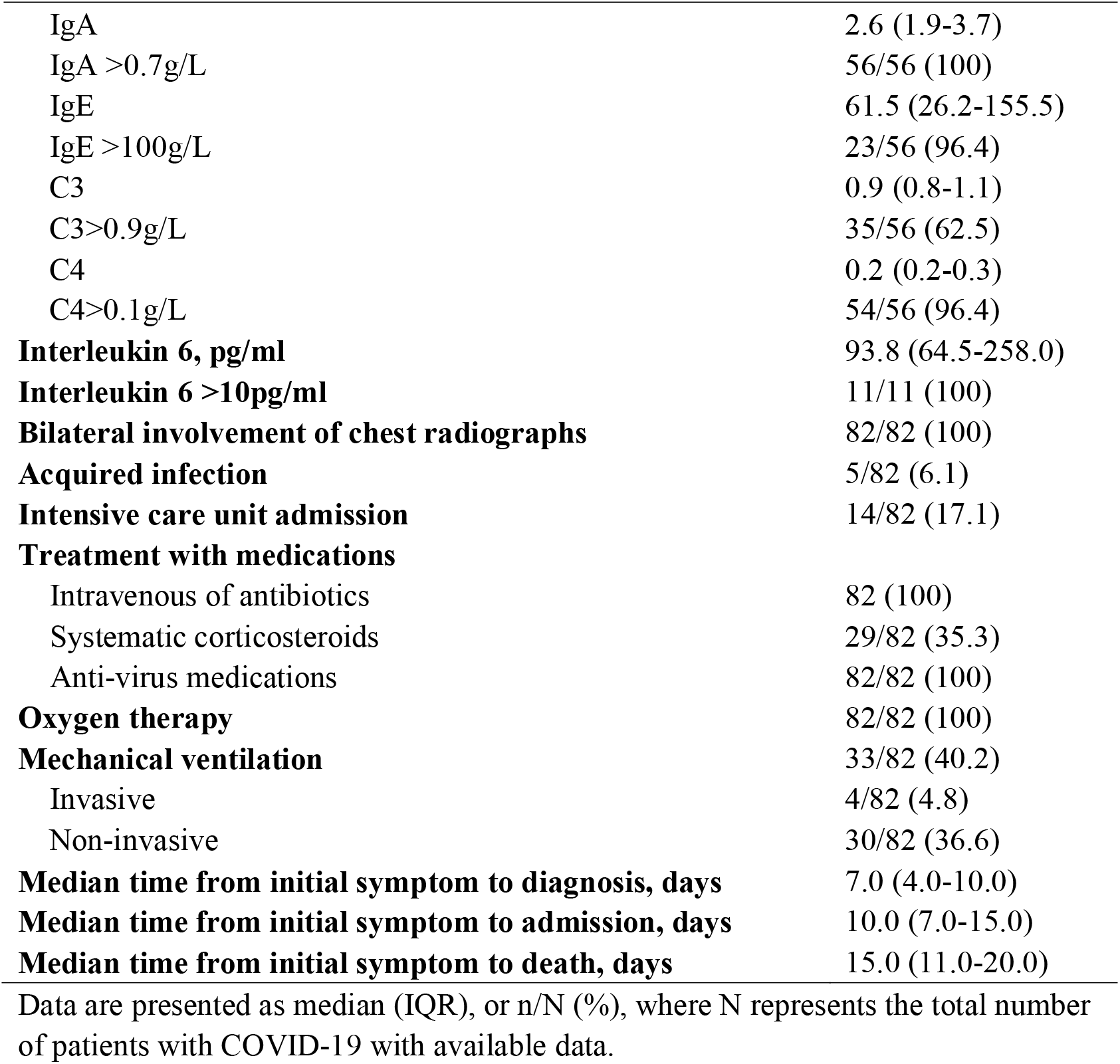
Initial symptoms, laboratory analyses, radiological findings, treatment, and survival time of dead patients with COVID-19

Furthermore, in the last 24 hours of the death, lymphopenia (73.7%), neutrophilia (100%), and thrombocytopenia (63.2%) were continuously present. Increased C-reactive protein level, high NLR, increased lactate dehydrogenase, and increased D-dimer were found in all patients. The incidence of neutrophilia was increased from 74.3% to 100%, and the incidence of lymphopenia was reduced from 89.2% to 73.7%. The incidence of high creatinine and blood urea nitrogen were increased from 15.3% to 45.0%, and 48.6% to 85.0%, respectively. High level of IL-6 (>10U/L) remained in all detected patients. More than half had a pH value of less than 7.35 (52.9%), and a pO_2_ value of less than 60 mmHg (70.6%) (table 4).

**Table 4.**
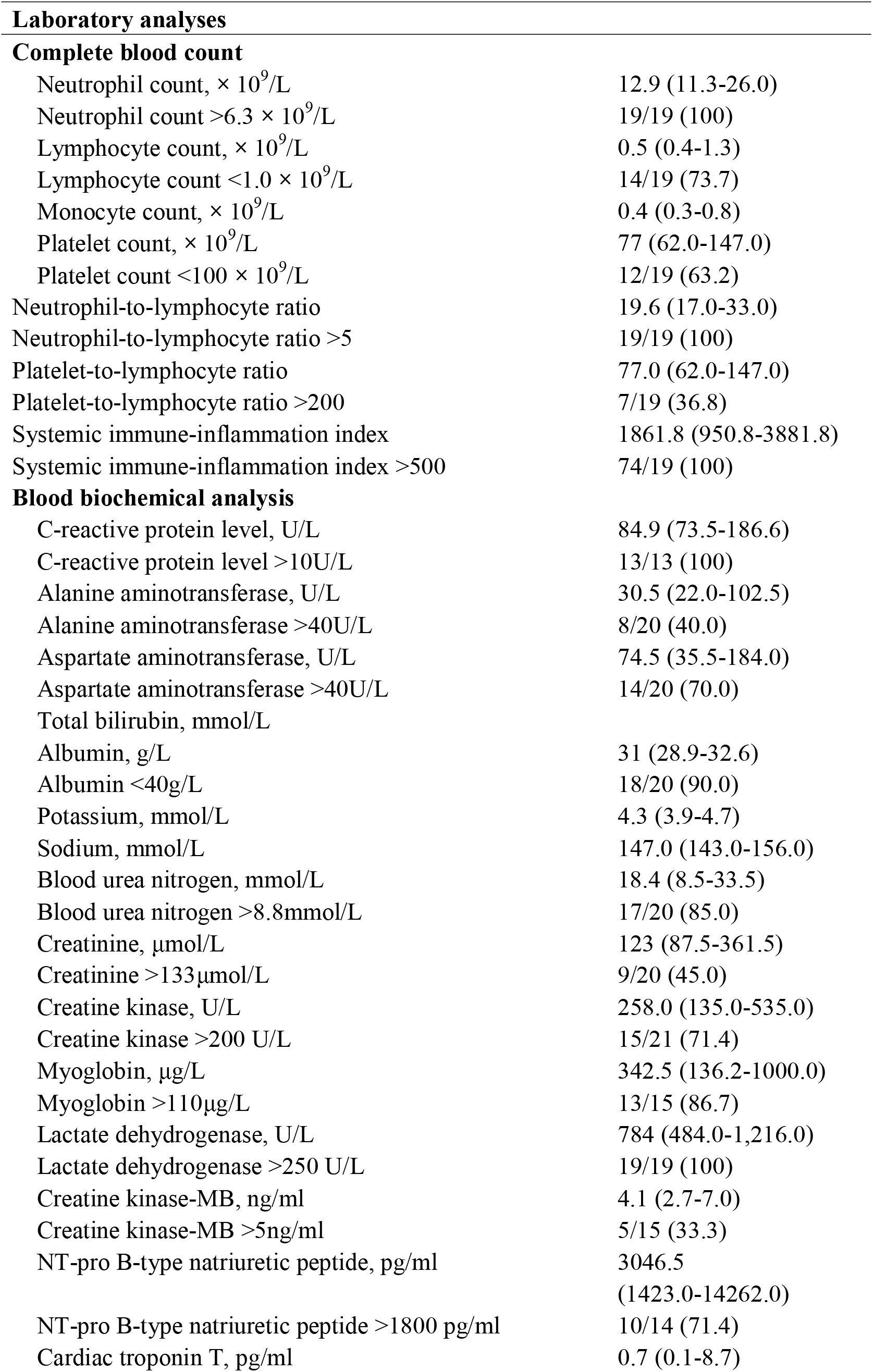
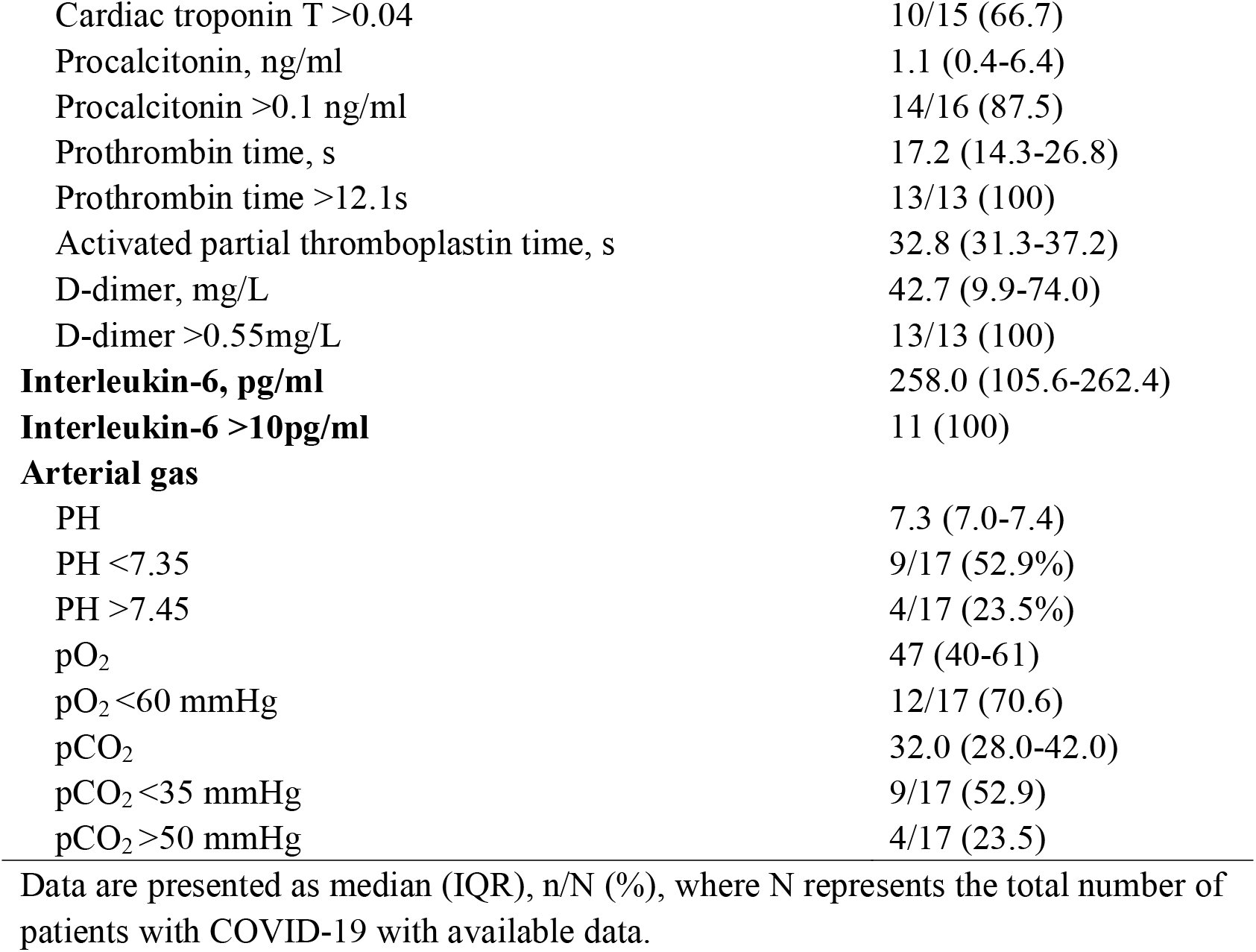
Laboratory analyses of dead patients in the last 24 hours of the death

|  |  |
| --- | --- |
| Neutrophil count, $\times 10^9/L$ | 12.9 (11.3-26.0) |
| Neutrophil count $>6.3 \times 10^9/L$ | 19/19 (100) |
| Lymphocyte count, $\times 10^9/L$ | 0.5 (0.4-1.3) |
| Lymphocyte count $<1.0 \times 10^9/L$ | 14/19 (73.7) |
| Monocyte count, $\times 10^9/L$ | 0.4 (0.3-0.8) |
| Platelet count, $\times 10^9/L$ | 77 (62.0-147.0) |
| Platelet count $<100 \times 10^9/L$ | 12/19 (63.2) |
| Neutrophil-to-lymphocyte ratio | 19.6 (17.0-33.0) |
| Neutrophil-to-lymphocyte ratio $>5$ | 19/19 (100) |
| Platelet-to-lymphocyte ratio | 77.0 (62.0-147.0) |
| Platelet-to-lymphocyte ratio $>200$ | 7/19 (36.8) |
| Systemic immune-inflammation index | 1861.8 (950.8-3881.8) |
| Systemic immune-inflammation index $>500$ | 74/19 (100) |
| **Blood biochemical analysis** |  |
| C-reactive protein level, U/L | 84.9 (73.5-186.6) |
| C-reactive protein level $>10U/L$ | 13/13 (100) |
| Alanine aminotransferase, U/L | 30.5 (22.0-102.5) |
| Alanine aminotransferase $>40U/L$ | 8/20 (40.0) |
| Aspartate aminotransferase, U/L | 74.5 (35.5-184.0) |
| Aspartate aminotransferase $>40U/L$ | 14/20 (70.0) |
| Total bilirubin, mmol/L |  |
| Albumin, g/L | 31 (28.9-32.6) |
| Albumin $<40g/L$ | 18/20 (90.0) |
| Potassium, mmol/L | 4.3 (3.9-4.7) |
| Sodium, mmol/L | 147.0 (143.0-156.0) |
| Blood urea nitrogen, mmol/L | 18.4 (8.5-33.5) |
| Blood urea nitrogen $>8.8mmol/L$ | 17/20 (85.0) |
| Creatinine, $\mu mol/L$ | 123 (87.5-361.5) |
| Creatinine $>133\mu mol/L$ | 9/20 (45.0) |
| Creatine kinase, U/L | 258.0 (135.0-535.0) |
| Creatine kinase $>200 U/L$ | 15/21 (71.4) |
| Myoglobin, $\mu g/L$ | 342.5 (136.2-1000.0) |
| Myoglobin $>110\mu g/L$ | 13/15 (86.7) |
| Lactate dehydrogenase, U/L | 784 (484.0-1,216.0) |
| Lactate dehydrogenase $>250 U/L$ | 19/19 (100) |
| Creatine kinase-MB, ng/ml | 4.1 (2.7-7.0) |
| Creatine kinase-MB $>5ng/ml$ | 5/15 (33.3) |
| NT-pro B-type natriuretic peptide, pg/ml | 3046.5<br>(1423.0-14262.0) |
| NT-pro B-type natriuretic peptide $>1800 pg/ml$ | 10/14 (71.4) |
| Cardiac troponin T, pg/ml | 0.7 (0.1-8.7) |
| Cardiac troponin T >0.04 | 10/15 (66.7) |
| Procalcitonin, ng/ml | 1.1 (0.4-6.4) |
| Procalcitonin >0.1 ng/ml | 14/16 (87.5) |
| Prothrombin time, s | 17.2 (14.3-26.8) |
| Prothrombin time >12.1s | 13/13 (100) |
| Activated partial thromboplastin time, s | 32.8 (31.3-37.2) |
| D-dimer, mg/L | 42.7 (9.9-74.0) |
| D-dimer >0.55mg/L | 13/13 (100) |
| **Interleukin-6, pg/ml** | 258.0 (105.6-262.4) |
| **Interleukin-6 >10pg/ml** | 11 (100) |
| **Arterial gas** |  |
| PH | 7.3 (7.0-7.4) |
| PH <7.35 | 9/17 (52.9%) |
| PH >7.45 | 4/17 (23.5%) |
| pO<sub>2</sub> | 47 (40-61) |
| pO<sub>2</sub> <60 mmHg | 12/17 (70.6) |
| pCO<sub>2</sub> | 32.0 (28.0-42.0) |
| pCO<sub>2</sub> <35 mmHg | 9/17 (52.9) |
| pCO<sub>2</sub> >50 mmHg | 4/17 (23.5) |

Data are presented as median (IQR), n/N (%), where N represents the total number of patients with COVID-19 with available data.

As shown in table 3, a total of 14 patients (17.1%) were treated in ICU. Patients received oxygen therapy (100%), mechanical ventilation (40.2%), including 4 invasive mechanical ventilation (4.8%). All patients received intravenous of antibiotics and anti-virus medications, and systematic corticosteroids were used in 29 patients (35.3%).

The median time from initial symptom to death was 15 days (IQR 15-20) and a significant association between aspartate aminotransferase (p=0.002), alanine aminotransferase (p=0.037) and time from initial symptom to death were interestingly observed (figure 1A-C).

**Figure 1:**
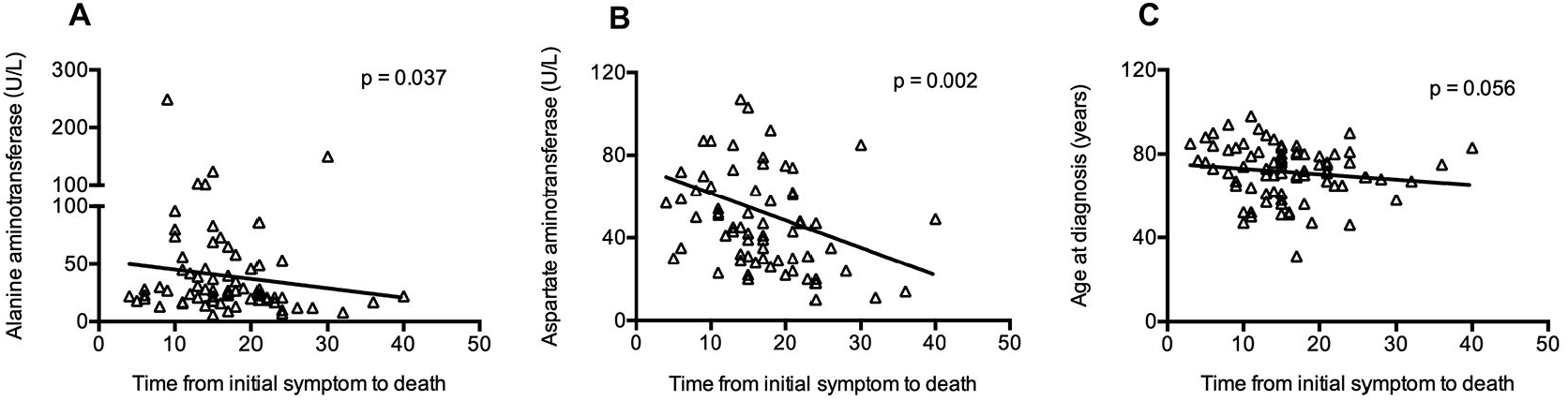
The association between clinical features and time from initial symptom to death. (A) Alanine aminotransferase (B) Aspartate aminotransferase (C) Age

## Discussion

To our knowledge, this is the first study to describe the clinical characteristics of dead patients with COVID-19. The mortality of 6.2% from current study was lower than that of SARS infection in 2003. However, the mortality rate from this center is a little bit higher than previously reported.^9^ We speculated the reason might be that fewer patients in our study were transferred to the ICU in time when their condition rapidly worsened. On the other hand, limited death cases, less than 15 patients were included in their cohort,^8,9^ while much more death cases were included in the present study.

Our study firstly focused on the epidemiological characteristics of dead patients with COVID-19. Several factors were responsible for the death of these patients included in this study. A majority of patients were older than 60 years in our study, and a borderline significant association between age and time from initial symptom to death were interestingly observed. These results are consistent with that older age was more likely occurred in critically ill patients.^16^ Moreover, we found underlying diseases such as hypertension, heart disease and diabetes were very common in our death cases, and 30.5% of patients had 2 or more comorbidities. These features are consistent with previous report that patients with underlying diseases more likely developed to severe illness.^9^ Cancer patients is comprised of 7.3% in our cohort, much higher than cancer morbidity, suggest that cancer patients more likely develop to severe disease, even death. These results are consistent with the findings from a national wide analysis in China.^17^ Immune deficiency to virus infection seems to be the common features in older males with comorbidities.

We further analyzed the cause of death case with COVID-19 and found that respiratory failure remained the leading cause of death. It has been reported that the binding receptor for SARS-CoV-2, ACE2 is mainly expressed in blood vessels and lung alveolar type II (AT2) epithelial cells,^13^ Similar to the SARS-CoV and MERS-CoV, SARS-CoV-2 can directly attack ACE2-expressing cells.^13^ Indeed, pathological findings indicated that infected lungs with SARS-CoV-2 present as ARDS, pulmonary edema with hyaline membrane formation, evident desquamation of pneumocytes.^18^ Therefore, our finding that respiratory failure is the leading cause of death, is consistent with the underlying pathological mechanism of COVID-19.

Besides respiratory failure, cardiac failure, hemorrhage, renal failure and even MOF were also recognized as the cause of death by COVID-19 in our study. Laboratory findings also revealed cardiac, hepatic, and renal damage in some of patients. We also observed a significant association between aspartate aminotransferase, alanine aminotransferase and time from initial symptom to death. These clinical phenomena could be explained by virus itself attack and cytokine release storm (CRS) mediated tissue damage. First, ACE2 expression is also found in the kidney, heart, and liver etc, therefore SARS-CoV-2 could invade the cells of above tissues, reproduce and damage these organ.^14,15^ Second, virus infection and subsequent tissue damage either in the lung or other target organ could elicit immune cells to produce pro-inflammatory cytokines, namely CRS, ultimately injury the tissue and cause target organ failure. Obviously, increased amounts of cytokines, including IL-1β, IL-6, and monocyte chemotactic protein-1 (MCP-1), are associated with severe lung injury in patients infected with SARS-CoV and MERS-CoV.^19,20^ A recent study showed high levels of IL-1β, interferon γ-induced protein 10, and MCP-1 occurred in serum of patients infected with SARS-CoV-2, which probably leaded to the activation of T-helper-1 cell response^1^. In the present report, high level of IL-6 of more than10U/L and C-reactive protein were detected in all patients, even in the last 24 hours prior to death.

We also depicted the immune status of COVID-19 patients with severe illness. Most of patients in our study presented as neutrophilia and lymphopenia on admission, specifically reduced CD3+, CD4+, and CD8+ T-cell counts were observed in some patients. A high NLR was also observed on the admission and 24 hours before the death. These results, consistent with the previous findings found in the patients with severe illness, suggest perturbation of immune system contribute the pathogenesis of SARS-CoV-2. These observations could be also explained why older males with comorbidities likely succumb to COVID-19.

Our study has some limitations. Firstly, some patients did not receive timely supportive interventions such as admission to ICU, because increasing number of severe patients occurred in a short period. However, present data could partially be scenario where COVID-19 patients progress in a natural pathophysiology rather than outcome from intervention by treatment. Secondly, consecutive detection of cytokines was lacking, which fail to truly monitor the severity of CRS. Thirdly, organ damage could originate from a history of medication including nonsteroidal anti-inflammatory drugs, antibiotics, and traditional Chinese medicine which are associated with renal or liver injury.^21,22^ In our study, all patients received intravenous of antibiotics and anti-virus drugs.

Overall, from the point of view of the causes of death, we presented the clinical characteristics of patients with COVID-19. Lung injury begins with an insult to the lung epithelium mainly attacked by SARS-CoV-2 itself because of ACE2 expressed in the lungs, which leads to most common respiratory failure. Other organs or tissues, more or less, are potentially damaged through direct attack from SARS-CoV-2. In addition, damages of multiple systems including the lungs, might originate with systemic damage due to CRS following SARS-CoV-2 infection. Considering the pandemic potential and moderate threaten of COVID-19 for population with multiple underlying diseases, further studies are required to focus on pathology and pathophysiology of tissue injury caused by SARS-CoV-2 infection, especially on the activation process of immune response and cytokines storm.

## Data Availability

The data that support the findings of this study are available from the corresponding author on reasonable request. Participant data without names and identifiers will be made available after approval from the corresponding author and National Health Commission. After publication of study findings, the data will be available for others to request. The research team will provide an email address for communication once the data are approved to be shared with others. The proposal with detailed description of study objectives and statistical analysis plan will be needed for evaluation of the reasonability to request for our data. The corresponding author will make a decision based on these materials. Additional materials may also be required during the process.

## Contributors

JW, BZ and QS had the idea for and designed the study and had full access to all data in the study and take responsibility for the integrity of the data and the accuracy of the data analysis. BiZh, YZ and YQ contributed to writing of the report. BZ contributed to critical revision of the report. JW and BiZh contributed to the statistical analysis. All authors contributed to data acquisition, data analysis, or data interpretation, and reviewed and approved the final version.

## Acknowledgments

We acknowledge all health-care workers involved in the diagnosis and treatment of patients in Eastern Campus, Renmin Hospital, Wuhan University; we thank Prof Hong Zhou and Jiang Zheng for guidance in manuscript preparation.

